# An international comparison of the second derivative of COVID-19 deaths after implementation of social distancing measures

**DOI:** 10.1101/2020.03.25.20041475

**Authors:** W. T. Pike, V. Saini

## Abstract

This work compares deaths for confirmed COVID-19 cases in China to eight other countries, Italy, Spain, France, USA, UK, Germany, Netherlands and South Korea. After implementing varying intensities and timing of social distancing measures, several appear to be converging onto the decline in the daily growth rate of deaths, or relative second derivative of total deaths, seen in China after the implementation an aggressive social distancing policy. By calculating future trajectories in these countries based on the observed Chinese fatality statistics, an estimate of the total deaths and maximum daily death rates over a defined period of time is made. Our lower bound estimate for the United Kingdom based on the real data approximates the lower bound estimate of the recent modelling study of Ferguson et al. [1]. These results suggest there may be a threshold of effective public health intervention. Our method of viewing the data may be helpful in monitoring the course of the epidemic, judging the effectiveness of implementation, and monitoring the relaxation of social distancing.

## Introduction

The global 2019-nCov pandemic currently underway has resulted in massive stresses on national health care systems, escalating deaths, and major disruptions in economic life in response to public health interventions designed to mitigate the pace of transmission. Modeling studies have shown a wide range of potential outcomes, depending on intensity of intervention. The trade-offs between the economic costs of preventive interventions and the health impacts of COVD19 have generated political controversy. As real-world data have begun to emerge, opportunities to illuminate these issues may be available.

## Methods

Data was obtained from European Centre for Disease Control website [2] on 24/03/2020. All reported deaths from confirmed cases of COVID19 were extracted into a time series of daily deaths for eight countries: China, Italy, Spain, France, USA, UK, Netherlands, Germany, and South Korea. In order to ensure that all included countries had sufficient counts to derive the necessary statistics, countries with fewer than 120 cumulative deaths were excluded. Daily fatality rates from the included countries were then used to calculate estimates of the relative second derivative of total deaths, *N*, 1/*N* d^2^*N*/d*t*^2^, for a period of at least ten days. All these countries have initiated social distancing measures at different points in time. Iran has been excluded as the very low variance in the data is not compatible with the statistics seen in the other seven countries analysed. The cumulative deaths for each country, *N*(*t*), were estimated by deriving a multiplier *N*_final_/*N*(*t*_conv_) at the time of convergence, *t*_conv_, to the Chinese trajectory, *N*_final_ being the total number of deaths in China and applying this multiplier to Chinese data for times beyond convergence. The daily deaths were derived by taking the differences in cumulative deaths.

## Results

Fig. 1 shows the relative second derivative plotted as a times series for each country compared to China, calculated after total deaths reached at least 20. An exponential rate corresponds to a constant value over time, but for China the daily rate has been falling from an initial 50% to very close to zero, with the fall occurring over a 30-day period after lockdown was announced for Wuhan on 23/01/2020. In fig. 1, the time series of the other countries have been aligned to best match the trajectory seen in China, with the announcement of lockdowns and implementation of social distancing measures in each country marked. Comparing to China, the daily growth rate in total deaths in Italy has been closely following the trajectory for the last 15 days subsequent to the announcement of lockdown in Lombardy on 8/3/2020.

**Figure 1:**
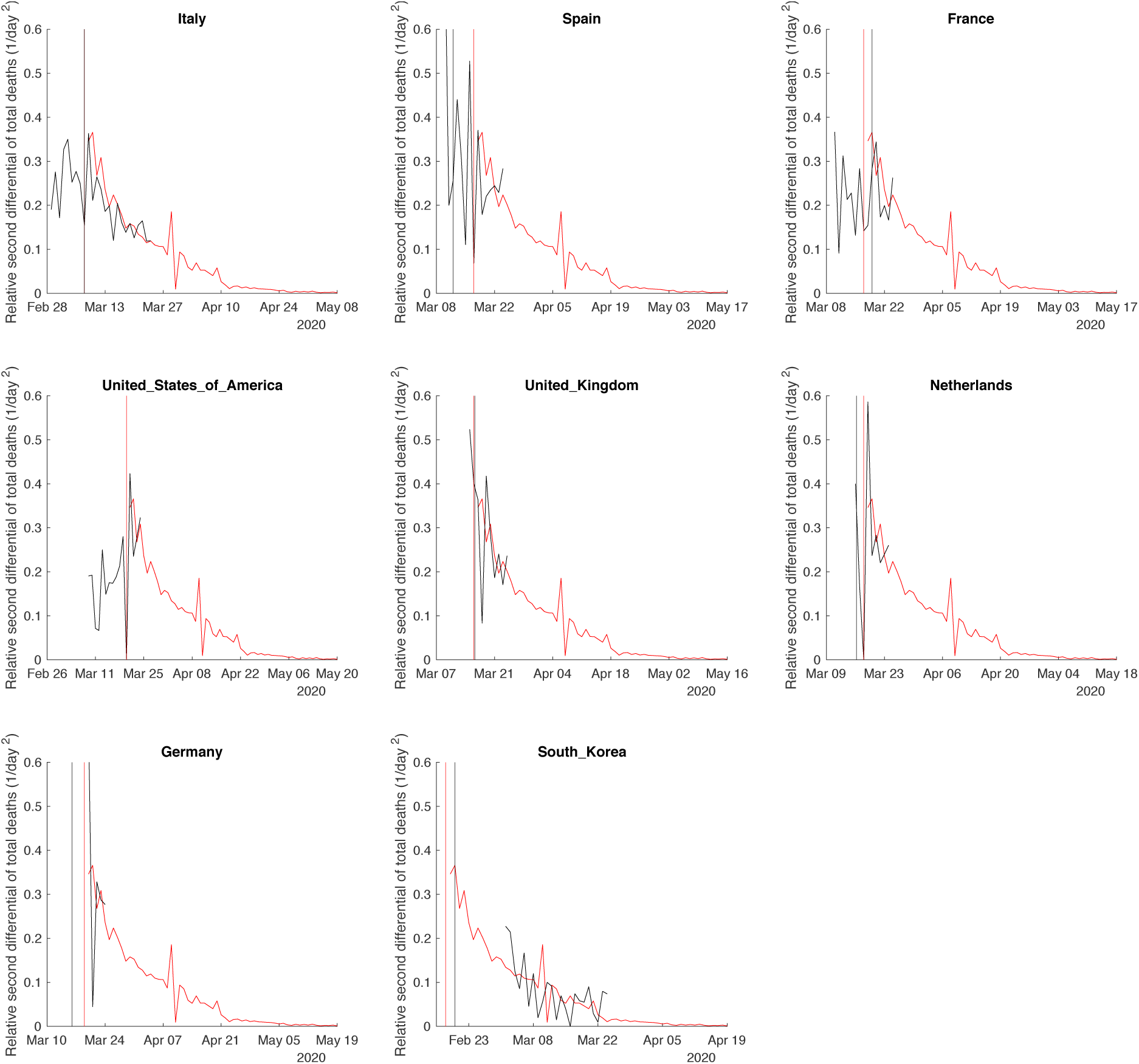
Relative second differential of total COVID-19-positive deaths in each country, plotted in black, up to 24/3/2020 compared to China, plotted in red. The time series have been shifted to best align with the trajectory seen in China after lockdown was announced in Wuhan on 23/01/2020, marked in red. The dates of social distancing for the comparator countries are shown in black. Data source: European Centre for Disease control, ECDC [1].

Spain has also seen some convergence on China’s trajectory for the last seven days following a national lockdown implemented on 12/3/2020, but with a higher variance. In the case of France, lock-down measures came into force on 16/3/2020, and the trajectory has converged, again with a higher variance, for the last six days. The USA also shows some convergence for the last four days, with lockdowns implemented only to the state level at the time of writing. The UK has converged on China’s trajectory for the last seven days, with a higher initial variance which in part is due to the sampling statistics from a relatively low fatality count to date; although no lockdown has been announced, general social distancing was advised by the government on 16/3/2020. Germany implemented social distancing in phases from Mar 8 until Mar 16 when stringent measures were announced nationally while the Netherlands did the same later and more slowly from 12/03 through 23/03/2020, ending in “targeted” lockdowns.

South Korea has seen a much lower rate of growth from the start and announced a number of suppression policies, including monitoring, testing and restricting non-essential travel from 20/2/2020. It converged on a later period of China’s trajectory, over a period of more than 20 days.

The multiplier curve for estimating total and daily deaths at the point of convergence is shown in Fig 2 and Fig. 3 shows the results using this multiplier if all of these countries continue to converge onto the Chinese trajectory. For example, on 4/3/2020 when South Korea converged onto the Chinese trajectory its total deaths were 32. The rate at that time corresponds to the rate seen in China on 8/2/2020, resulting in a corresponding multiplier of 4.5, and predicted total deaths of 144. Current cumulative deaths in South Korea are 103, consistent with South Korea continuing to follow China’s trajectory, with an estimate, on 24/03/2020, of 130 total deaths, within 10% of the initial estimate. The conditions for a reduced death toll according to this calculation are a low number of deaths prior to convergence, and a point of convergence that is at an advanced point in the Chinese trajectory. South Korea fulfils both conditions. As this multiplier depends on the date of convergence, the sensitivity of this estimate can be calculated for an offset of ±1 day. On the basis of this assumption, the projected total deaths and a sensitivity range are shown in Table 1.

**Table 1:** Estimates (actual for China for 20/03/2020) of the total COVID-19 deaths and the maximum daily deaths (3-day average) based on the listed countries following the growth trajectory of China from 20/03/2020. Lower and upper sensitivities are based on an offset from the current best match to China’s trajectory of plus or minus one day. The date of maximum daily deaths corresponds to the estimated value.

|  | Total deaths |  |  | Maximum daily deaths |  |  |  |
| --- | --- | --- | --- | --- | --- | --- | --- |
|  | Estimate | Lower sensitivity | Upper sensitivity | Estimate | Lower sensitivity | Upper sensitivity | Date |
| China | 3280 | Actual | to date | 150 | Actual | to date | 23/02/2020 |
| Italy | 28000 | 25000 | 32000 | 1300 | 1200 | 1500 | 27/03/2020 |
| Spain | 46000 | 37000 | 60000 | 2200 | 1700 | 2800 | 04/04/2020 |
| France | 18000 | 14000 | 23000 | 800 | 660 | 1100 | 08/04/2020 |
| USA | 28000 | 22000 | 41000 | 1300 | 1000 | 1900 | 08/04/2020 |
| UK | 5700 | 4700 | 7100 | 260 | 210 | 330 | 05/04/2020 |
| Netherlands | 6000 | 4700 | 7500 | 280 | 220 | 350 | 24/03/2020 |
| Germany | 4000 | 3200 | 5300 | 190 | 150 | 250 | 09/03/2020 |
| South Korea | 150 | 140 | 160 | 2 | 2 | 2 | 06/04/2020 |

**Figure 2:**
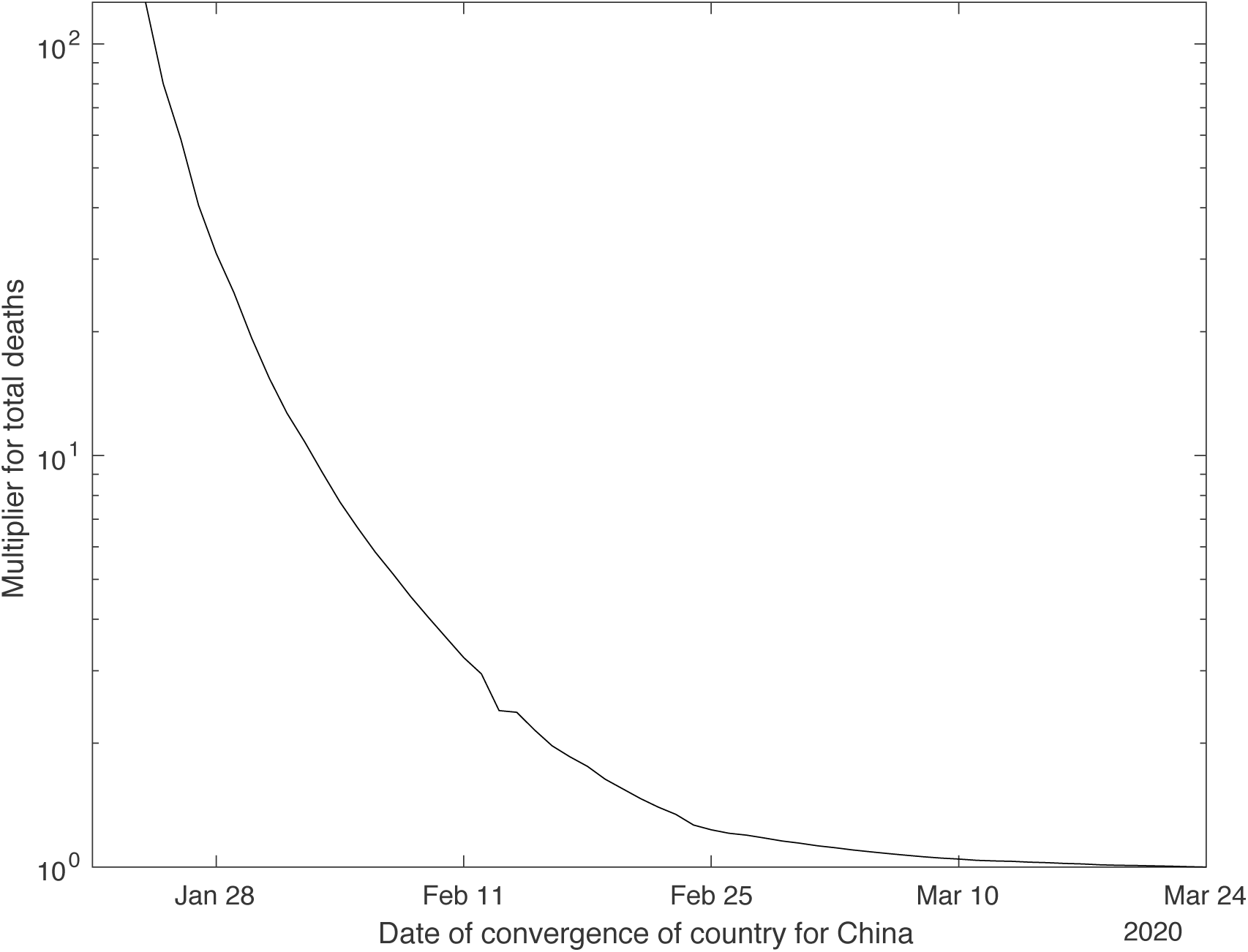
Multiplier for current total deaths for a country outside China to give the final total deaths, plotted as a function of the date of convergence for the Chinese dataset.

**Figure 3:**
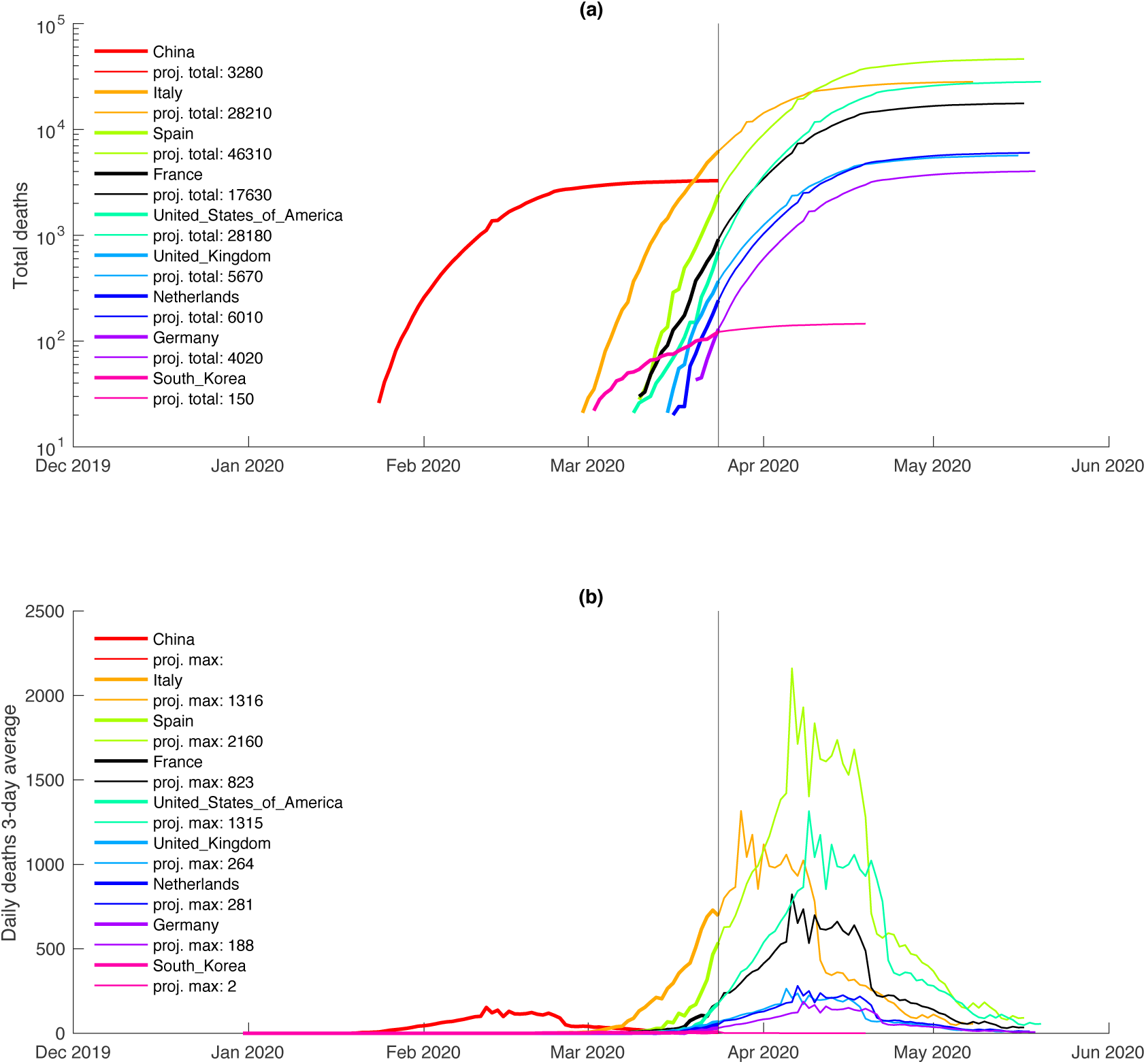
Estimates of (a) the total COVID-19 deaths and (b) daily deaths for the listed countries derived by following the trajectory of China from 24/03/2020.

## Discussion

The observed convergences to date, at an early point in the Chinese trajectory, for Italy, Spain, France, USA, UK, Netherlands and Germany, would indicate convergence of the combined transmission and fatality rate for these eight countries. If a similar fatality rate is assumed, this would imply these countries are converging on the same trajectory in transmission rates, the stated goal of efforts at social distancing.

Although the strength of social distancing has varied in all eight countries, with China’s the most restrictive and closely monitored, overall these measures appear to be sufficient at this time to produce a similar fall in cumulative death rates. This implies there may be a threshold of intervention sufficient to achieve the desired trajectory. South Korea’s policies, with intervention at an early stage, has been successful in reducing the rate of increase in deaths to that seen in the latter stages of lockdown in Wuhan. If the present trend continues, the Netherlands’ policy of “targeted” lockdowns may also be achieving a similar result.

These estimates are based on each of the eight countries following China’s trajectory from the present. Due to their deaths before convergence, most would still expect from two to eight times the number of total and daily deaths. Based on this model, it would be unexceptional to see daily deaths of 1000 in Italy in the near future even as it remains on the trajectory, despite following the downwards trajectory of growth in daily death rates seen in China. The exceptions are South Korea, which is much lower, and Germany and the Netherlands which appear to be roughly equivalent.

In general, this analysis suggests that early adoption of social distancing is more effective than delayed implementation, even of highly restrictive measures. If China’s policies aimed at achieving suppression represent a maximum possible reduction in transmission from initially high rates, then estimates based on their curve will undercount both the total deaths and maximum death rates for other countries and hence represent a reasonable lower bound for modelling: the lowest estimate of total deaths in the models in Ferguson et al. [2] is 5,600, similar to the expected value of 5,700 we predict if the UK follows China’s trajectory from 24/03/2020.

## Conclusions

Our analysis suggests there may be a threshold of public health intervention beyond which a decline in death rates begins to occur. This simple method of viewing the data may be helpful in monitoring the course of the epidemic at national and regional levels, judging the effectiveness of implementation in individual countries, planning for and monitoring the relaxation of social distancing, and potentially improving time estimates for economic projections.

## Data Availability

Data available at: https://www.ecdc.europa.eu/en/publications-data/download-todays-data-geographic-distribution-covid-19-cases-worldwide

## References

[1] Ferguson et al., Impact of non-pharmaceutical interventions (NPIs) to reduce COVID-19 mortality and healthcare demand https://www.imperial.ac.uk/media/imperial-college/medicine/sph/ide/gida-fellowships/Imperial-College-COVID19-NPI-modelling-16-03-2020.pdf

[2] https://www.ecdc.europa.eu/en/publications-data/download-todays-data-geographic-distribution-covid-19-cases-worldwide

